# Forecasting the COVID-19 Pandemic: Lessons learned and future directions

**DOI:** 10.1101/2021.11.06.21266007

**Authors:** Saketh Sundar, Patrick Schwab, Jade Z.H. Tan, Santiago Romero-Brufau, Leo Anthony Celi, Dechen Wangmo, Nicolás Della Penna

## Abstract

The Coronavirus Disease 2019 (COVID-19) has demonstrated that accurate forecasts of infection and mortality rates are essential for informing healthcare resource allocation, designing countermeasures, implementing public health policies, and increasing public awareness. However, there exist a multitude of modeling methodologies, and their relative performances in accurately forecasting pandemic dynamics are not currently comprehensively understood.

In this paper, we introduce the non-mechanistic MIT-LCP forecasting model, and assess and compare its performance to various mechanistic and non-mechanistic models that have been proposed for forecasting COVID-19 dynamics. We performed a comprehensive experimental evaluation which covered the time period of November 2020 to April 2021, in order to determine the relative performances of MIT-LCP and seven other forecasting models from the United States’ Centers for Disease Control and Prevention (CDC) Forecast Hub.

Our results show that there exist forecasting scenarios well-suited to both mechanistic and non-mechanistic models, with mechanistic models being particularly performant for forecasts that are further in the future when recent data may not be as informative, and non-mechanistic models being more effective with shorter prediction horizons when recent representative data is available. Improving our understanding of which forecasting approaches are more reliable, and in which forecasting scenarios, can assist effective pandemic preparation and management.

## II. Introduction

During the COVID-19 pandemic, the Centers for Disease Control and Prevention (CDC) broadcasted an open call to forecast COVID-19 cases and deaths at a state level. All data scientists and research teams with models that predicted the course of the COVID-19 pandemic were welcomed to submit their models to the CDC COVID-19 Forecast Hub. CDC COVID-19 Forecast Hub pipeline teams were invited to submit predictions of the numbers of new cases, hospitalizations, and deaths in future days, weeks, and months, for the county, state, and national levels in the U.S.

Groups across the U.S. assembled organically and a community emerged that consisted of individuals with different backgrounds and expertise but with a shared vision of forecasting to inform policy. Despite the existence prior to COVID-19 of troves of data which could be analyzed for real-time prediction in the event of a disaster, the pandemic exposed the fault lines in our data-verse. The world was ill-prepared to leverage the data tsunami that no one group, university, or country had the ability to harness in order to understand and forecast the trajectory of the pandemic.

Accurate predictions concerning future disease spread, mortality, and analysis of what-if scenarios can be critical for guiding decision-makers in a pandemic. Government officials may utilize this information to issue public health guidelines and policy, while health system leadership may utilize it to make resource allocation and triage decisions. Decisions can be characterized as upstream or downstream of transmission drivers. Certain upstream decisions, such as mandatory lockdowns, affect the spread of infection. Other downstream decisions, such as canceling elective surgeries in order to free up hospital beds to treat infected patients, are unlikely to meaningfully affect the spread of infection.

Forecasting models typically fall along a spectrum of what we could call ‘mechanistic’ to ‘non-mechanistic’ models. Mechanistic models incorporate a given understanding of the underlying causal structure of the data generation process. Non-mechanistic models do not incorporate such structural assumptions.

Mechanistic models are necessary for studying the potential effect of decisions that interact with underlying disease spread dynamics since this is an inherently counterfactual task. Mechanistic models allow the data of the observed portion of the action space to be projected into estimates in the unobserved parts.

While evaluating the accuracy of these forecasts in the unobserved actions is not a feasible empirical exercise, it stands to reason that if the structure of the data generation process is being captured, it should not make worse predictions in the observed part of the action space than models that do not make such assumptions.

Our approach utilized standard machine learning techniques that impose little structure on the data distribution, while other groups employed a much more mechanistic modeling approach. Our empirical results provide a benchmark against which to measure mechanistic models, and thus to evaluate how the implied structure they impose performs in those parts of the action space that are observed.

This work presents a non-mechanistic approach to real-time forecasting of U.S. COVID-19 mortality using a gradient-boosted regressor model (named MIT-LCP).

## III. Related Work

### A. Non-mechanistic approaches to forecasting

Non-mechanistic approaches to forecasting have been implemented in numerous application domains from influenza forecasting [11] to population dynamics of beetles [15]. In prior epidemics such as the Ebola epidemic, flexible non-mechanistic or semi-mechanistic models have been implemented since parameterizing mechanistic models was often difficult in real-time, when information on behavioral changes, interventions and routes of transmission were not readily available [12]. In both the influenza endemic and Ebola epidemic, non-mechanistic models had promising results in real-time forecasting when compared to traditional statistical models.

### B. CDC COVID-19 Forecast Hub

The CDC COVID-19 Forecast Hub has featured 50+ models of both the non-mechanistic and mechanistic types [20]. In the mechanistic variety there are time dependent Susceptible-Exposed-Infectious-Removed (SEIR) models [25] which is a compartmental model [21]. There are also metapopulation models which are compartmental models that investigate interactions and movements among different subpopulations [22]. Some models are modified SEIR models, such as the Texas Tech [24] or Delphi [23] models which incorporate additional compartments for undetected cases and quarantined populations. Mechanistic models can also be combined with machine learning methods as is seen with the UMass model [8]. There were a number of non-mechanistic models on the CDC hub as well [26] - with a majority being regression models with augmentations such as the UMich model.

### C. Prior comparisons with non-mechanistic and mechanistic models

Machine learning and mechanistic models have been compared in multiple public health-adjacent contexts [27] from a biological context [28] to a clinical context [29]. These studies do not consider mechanistic and non-mechanistic models as direct competitors [28]. In the biological context, it was found that integrating non-mechanistic and mechanistic models yielded better performance than utilizing just one [28]. In the clinical setting, there exist studies that report on mechanistic models outperforming non-mechanistic machine learning models in breast cancer metastatic relapse prediction [29]. However, in particular in the context of COVID-19, there exist published examples of performant non-mechanistic models addressing clinical prediction tasks [30, 31]. To the best of our knowledge, there do not yet exist studies directly comparing mechanistic and non-mechanistic models in the public health setting.

## IV. Methods

### A. Data Sources

The MIT-LCP model is implemented using a gradient boosted regressor to forecast COVID-19 deaths at the state and national levels. It uses novel digital data sources including prior COVID-19 cases and deaths, demographic, socioeconomic, mobility, and healthcare-related county-level covariates for feature generation. The details and characteristics of the data sources are described in Table I. Table I lists the description, scope, time frame, and the number of features used for each dataset.

**TABLE I.** Data Sources for MIT-LCP Model.

| **Data Sources for MIT-LCP Model** |  |  |  |  |
| --- | --- | --- | --- | --- |
| *Data source* | *Description* | *Scope* | *Time frame* | *Number of Features* |
| JHU [1] | Daily US COVID-19 Deaths and Cases | County Level | 2020-2021 | 6 |
| COVID Tracking Project | Daily COVID-19 Tests | State Level | 2020-2021 | 3 |
| County-level Socioeconomic Dataset [2] | 300 variables that summarize population estimates, demographics, ethnicity, housing, education, employment and income, and healthcare system-related metrics | County Level | Static, data from 2018-2020 | 57 |
| PlaceIQ [3] | Exposure indices derived from PlaceIQ movement data | County Level | 2020-2021 | 2 |
| Hospitalizations | COVID-19 Hospital capacity and utilization from HHS | State Level | 2020-2021 | 2 |

In addition to serving as the target data for forecasts, the Johns Hopkins University Center for Systems Science and Engineering (JHU CSSE) deaths and cases data were an important component in the training of the model. 3-week lagged cases and deaths were used to predict 1-4 weeks ahead. The COVID-19 tracking project’s state-level testing data also served as a useful feature for forecasting deaths. The test positivity rate in a specific state was a good indicator of the severity of the pandemic at that time [4]. The county-level socioeconomic dataset was the only dataset used that was not in a time-series format. Although it was static, the variables in the dataset, specifically the demographics and education variables were valuable because certain groups of people are more vulnerable to COVID-19 [5], and the model can leverage these variables to predict mortality. Additionally, the healthcare variables from this dataset served as a strong indicator of each county’s healthcare system’s relative strength [6]. Previous studies have established that an increase in mobility metrics leads to an increase in COVID-19 cases and deaths [7], so PlaceIQ exposure indices were used in the feature set for training as well. State-level hospitalization data as correlates for pandemic severity were the final variable added to the feature set.

### B. Forecasted Data Targets

In accordance with the structure of the CDC Forecast Hub, we made probabilistic forecasts for 1–4 weeks ahead of incident and cumulative deaths for each county in the US, which were then aggregated to the state and national level.

The primary modeled quantity of the model was weekly incident deaths, which were added to previous forecasts for incident deaths as well as previous cumulative deaths to create the cumulative death forecast. The forecasts evaluated in this paper are state and national-level real-time weekly incident forecasts only. This means that they submitted forecasts weekly during the period of evaluation as new data arrived.

### C. Model Pipeline Architecture

Figure 1 shows the various stages of the MIT-LCP model pipeline. The first step in the forecasting pipeline was conducting data source retrieval for data sources that refreshed on a daily basis. Upon retrieval, the data were parsed and preprocessed, and used to generate specific features for the model. The datasets were then combined to generate additional features. The regression model was trained on the features and the strength of each feature was determined using SHAP (SHapley Additive exPlanations) values (Figure 2) [10].

**Figure 1:**
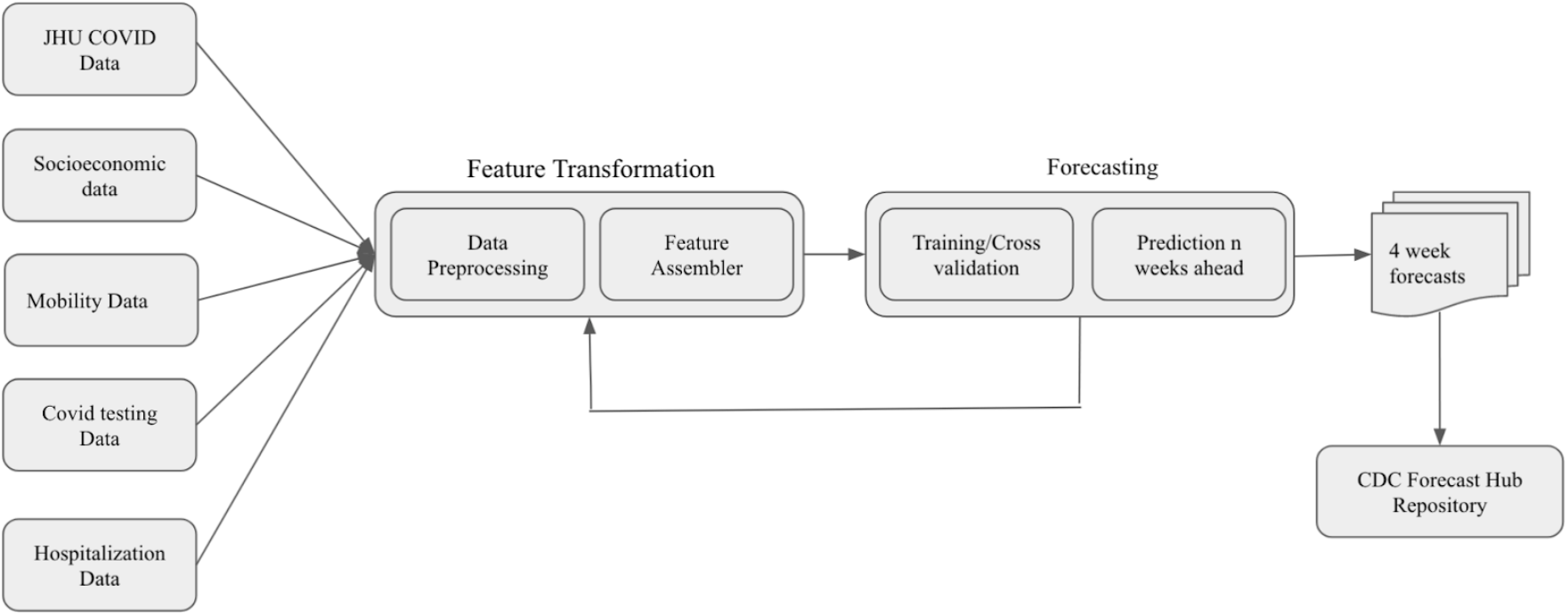
MIT-LCP Model Pipeline Architecture. Data are processed, and features are assembled. Training of the model occurs and features are reassembled. Afterwards predictions n weeks ahead are made and then submitted to the CDC forecast hub.

**Figure 2:**
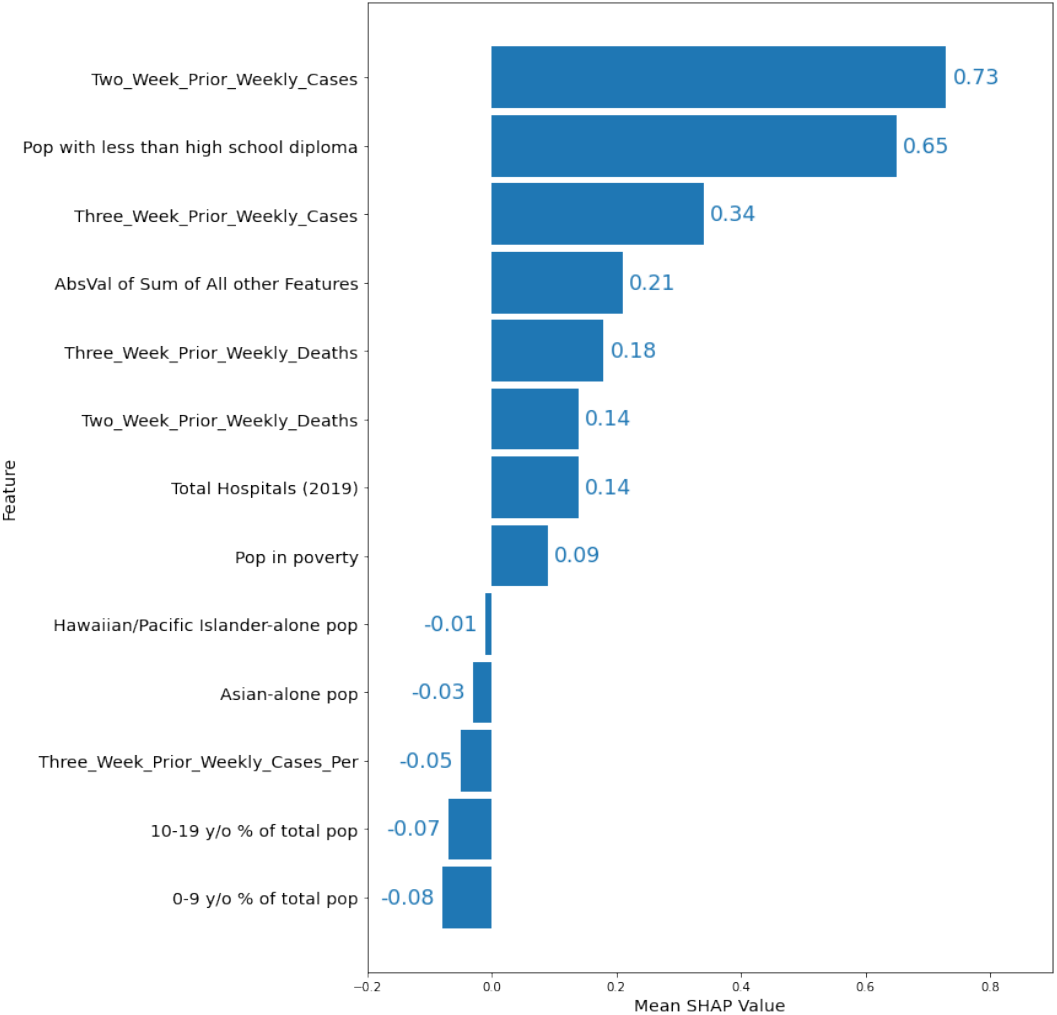
Mean absolute value of feature SHAP values

The features with higher relative strength were then separated from the rest, and the model was then trained again specifically on these features. Finally, the model predicted incident and cumulative deaths for 1-4 weeks ahead at the county-level for all the counties in the US, and these predictions were then aggregated to the state and national level. The forecasts were formatted for submission to the CDC Forecast Hub and then submitted to their Github repository.

### D. Software and Algorithms

The real-time forecasting model was implemented using Python 3.6 in Jupyter notebooks. Python packages used include NumPy [16], Pandas [17], scikit-learn [18], and XGBoost [9]. The model consisted of an ensemble of extreme gradient boosted trees of linear regressors trained with the mean squared error loss function. Gradient boosting involves iterative combinations of ensembles of weak prediction models into one strong learner. The package XGBoost uses second-order Taylor series to approximate the value of the loss function and reduces the likelihood of overfitting [9]. The model utilized hyper parameter optimization, the approach being a cross-validated grid search. The optimized hyper parameters were the learning rate, number of trees, and maximum tree depth. Since non-stationary time series data, like COVID-19 deaths or cases, can be problematic for cross validation, we used scikit-learn’s TimeSeriesSplit for training and validation instead of a k-fold split.

Among the most important features for the MIT-LCP model were the “Two Week prior Cases” and “Three Week prior Cases” in the area, which aligns with the typical period of time from infection to death of a COVID-19 patient. Additionally, the “Three Week prior Deaths” and “Two Week prior Deaths” also had a high SHAP value, as the model used past weeks’ deaths to predict the trend for the future weeks’ deaths. Another feature with a high SHAP value was the “Population with Less than a High School Diploma”, which might indicate hesitancy towards mask-wearing and vaccination in counties with larger populations of that category. “Population in Poverty” also had a high SHAP value, as did “Total Hospitals”, which line up with findings of past studies into the levels of transmission in areas with high poverty or areas with less access to healthcare. Features with the largest negative SHAP values did not have very significant impact compared to the features with the largest positive SHAP values - as they all have SHAP values less than -0.1

### E. Evaluation Metrics

The following three metrics were used for the evaluation of the forecasts. For the model training, the R2 Score was used. For comparison of the models, two evaluation metrics were used: FAPE and MAE.

- R2 Score - Coefficient of Determination was used in the training of the model, the objective in the training was to maximize the R2 score.
- FAPE - Forecast Absolute Percentage Error was used for the national level comparison. The metric was calculated using the actual value and the forecasted value of deaths for a given week.

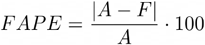
- MAE - Mean Absolute Error was used for the state-level comparison. The metric was calculated using actual values and the forecasted value of deaths for each state in a given week.

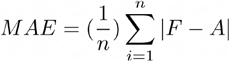

## V. Results

To evaluate the MIT-LCP model performance, seven other CDC Forecast Hub models were chosen for comparison. The mechanistic SEIR (Susceptible, Exposed, Infected, Resistant) models used for comparison were CovidAnalytics-DELPHI (Delphi), CU-select (Columbia), TTU-Squider (Texas Tech), JHU-APL, and UCLA-SuEIR (UCLA). UMich-RidgeTfReg (UMich) and UMass-MechBayes (UMass) [8] were among the best-performing models on the CDC Forecast Hub during this time period and they were also included for comparison. UMich is a non-mechanistic model, while UMass is a mechanistic model implemented using machine learning methods.

Figure 3 shows the national mortality Forecast Absolute Percentage Error (FAPE) over the course of 22 evaluation weeks. MIT-LCP had a lower median FAPE compared to 4 of 5 mechanistic SEIR models submitted to the Forecast Hub and similar or higher FAPE compared to UMich and UMass models. At the national level, the MIT-LCP model achieved a median FAPE of 15.05% across the 22 weeks for all forecast dates whereas the SEIR models except for the UCLA model had FAPEs in the range of 20-25% during the same period. MIT-LCP model also had a smaller interquartile range within the distribution of FAPEs compared to every mechanistic model in the comparison set except for the UMass model.

**Figure 3.**
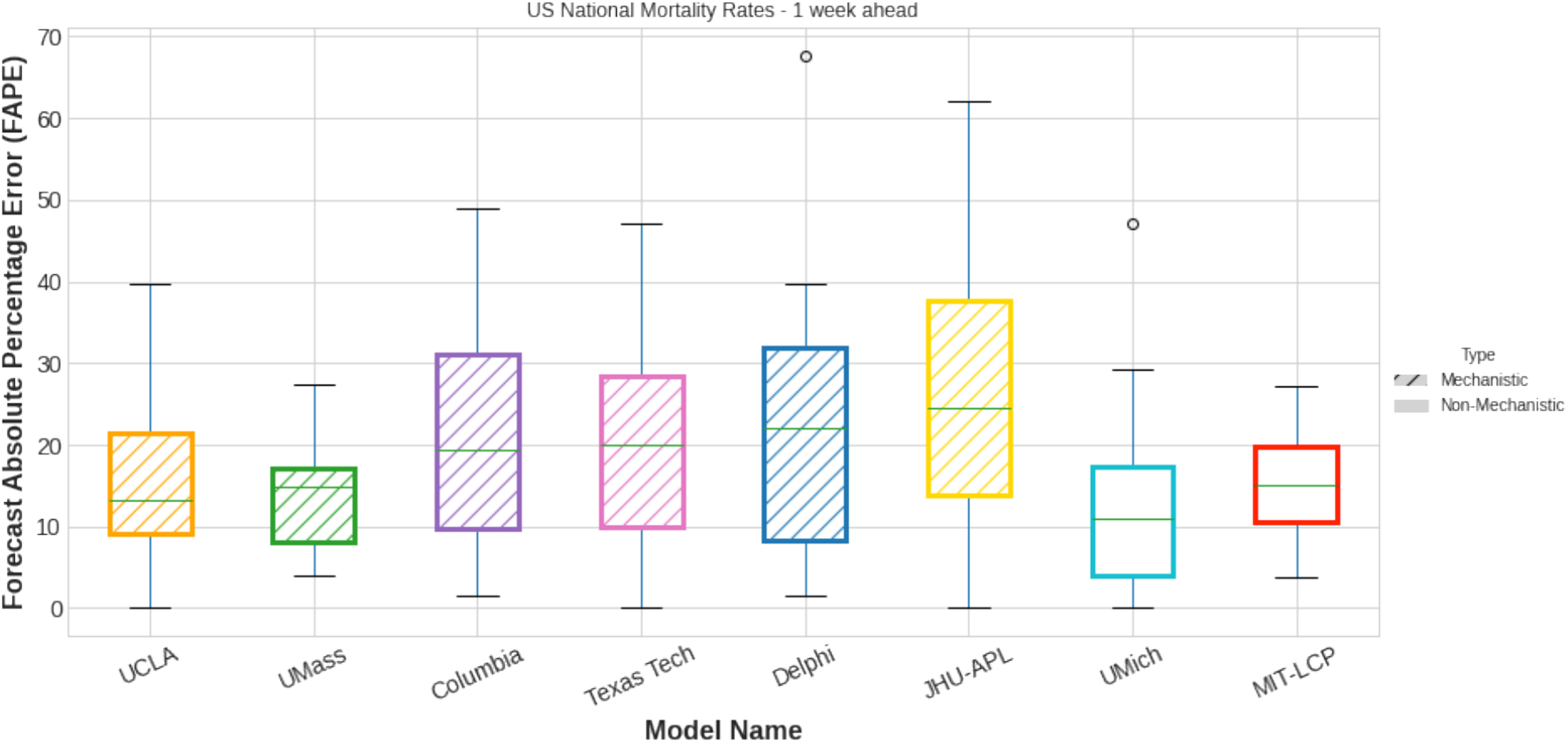
Comparison of MIT-LCP versus other CDC Forecast Hub Models in terms of Forecast Absolute Percentage Error (FAPE) for predicting national mortality rates 1 week in advance. Data covers the period from November 1^st^ 2020 to April 2^nd^ 2021.

Figure 4 shows US states mortality forecast Mean Absolute Error (MAE) for 1-4 week ahead predictions over the course of 22 weeks. MIT-LCP had a lower median MAE compared to most mechanistic models for the 1 week ahead target. MIT-LCP improved uniformly over most models for every target with MAE ranging 34-400 deaths across the 1–4 week ahead predictions. Only the UMass model had a lower distribution of errors than MIT-LCP across the 1, 2, 3, and 4 weeks ahead forecasts. Significant variability in the distribution of errors by forecast date for different target weeks reflects the difficulty in forecasting throughout the pandemic.

**Figure 4.**
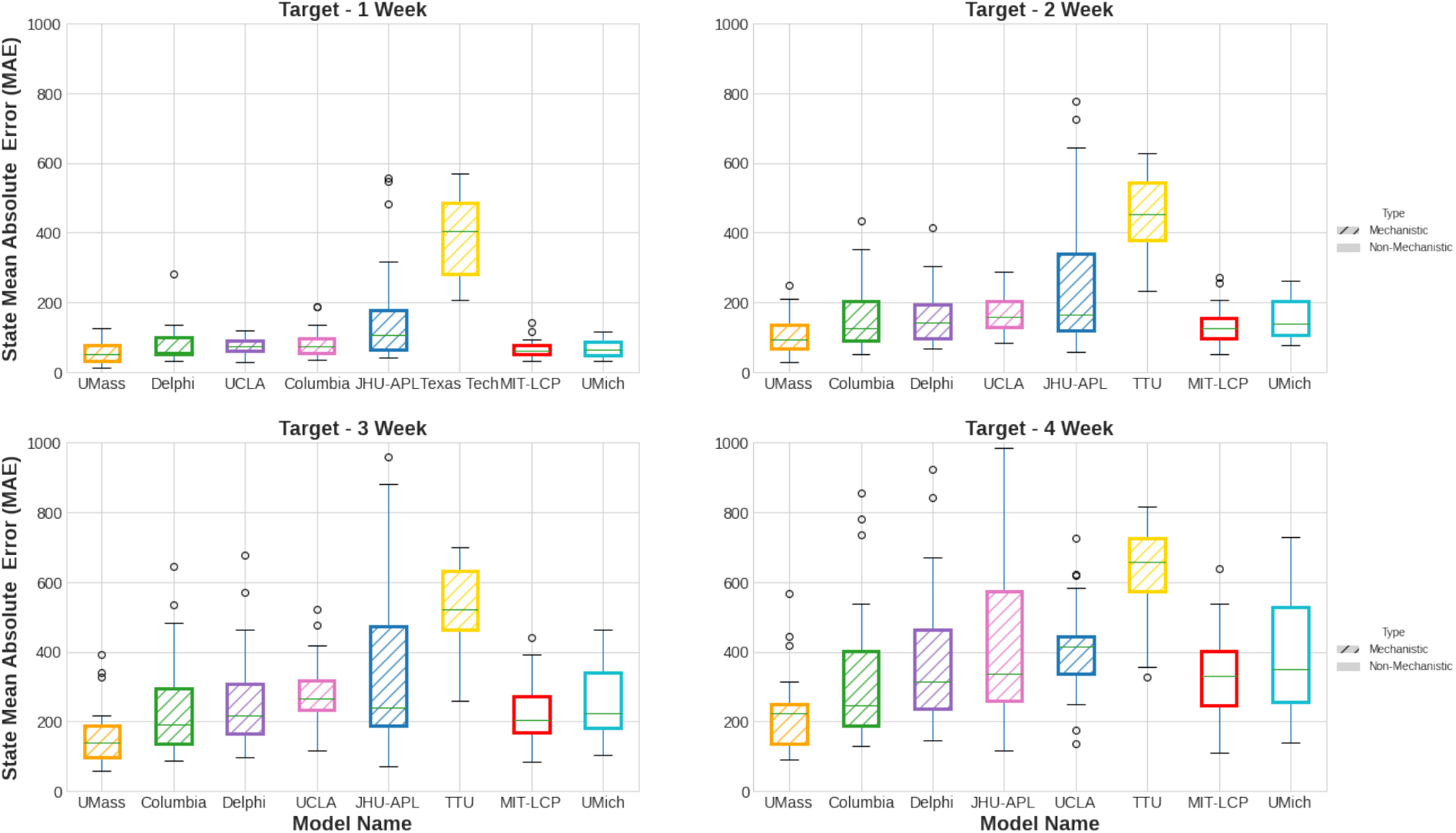
Comparison of MIT-LCP versus other CDC Forecast Hub Models in terms of Mean Absolute Error (MAE) for predicting US states’ mortality rates 1, 2, 3, and 4 weeks in advance. Data covers the period from November 1^st^ 2020 to April 2^nd^ 2021.

## VI. Discussion

Policymaking is especially challenging during an emerging infectious disease pandemic. Public health interventions and policies that curtail individual freedom, such as movement restrictions or isolation and quarantine orders, can be particularly challenging for policymakers in positions that are sensitive to public opinion and the prevailing political winds. Decision-makers nevertheless rely heavily on public health experts to advise them and provide compelling data that policymakers can utilize to defend their decisions to the public.

Accurate non-mechanistic forecasting models offer a critical tool for public health policy and practice to inform downstream policy decisions and facilitate -- particularly with regard to future pandemics that involve new or emerging diseases where sufficient knowledge to effectively inform mechanistic forecasting models is lacking.

For the COVID-19 pandemic, many mechanistic models underperformed compared to a non-mechanistic baseline. Mechanistic models that implemented the SEIR framework typically underperformed relative to other models. However, some mechanistic models utilized the SEIR framework with a more Bayesian approach -- such as the UMass-MechBayes model. This particular model performed quite well compared to both the mechanistic SEIR models and MIT-LCP’s non-mechanistic model.

One major limitation of many non-mechanistic models is the lack of causal inference. Integration of causal inference methods with machine learning in non-mechanistic models could improve the overall performance of the model. A second limitation is the lack of policy data integration with forecasting models. Datasets of policies and public health guidelines issued at the state and local-level could aid in the creation and advancement of forecasting models. With these policy datasets, conditional predictions could be standardized, where forecasts are generated depending on the policies that could be implemented. This improvement could increase the accuracy of forecasts as well as expand the potential impact of the forecasts on policy decisions.

## VII. Conclusion

Infectious disease forecasts provide critical data for informing public health policy and interventions. Mechanistic and non-mechanistic disease transmission forecasting models each have their own respective use case advantages and disadvantages, which can be used to complement the other. In terms of performance metrics, non-mechanistic forecasting models perform at least equally well as mechanistic models, even outperforming mechanistic models in some cases, and should be used in conjunction with each other. Utilizing both types of forecasting models and implementing improvement measures that are widely used in real-time forecasting can assist in more effectively preparing for future pandemics.

Finally, this paper calls attention to the community that organically grew around a shared purpose of leveraging existing data to forecast the trajectory of the pandemic in order to inform policy. This community spanned multiple institutions, disciplines, and expertise levels, drawing from academia, industry, and government, and has attracted contributors from senior investigators all the way to high school students. This paper’s group of authors reflects that diverse, cross-disciplinary community. Prior to COVID-19, the authors had formed a global consortium called MIT Critical Data to advance health data science, which consisted of healthcare practitioners, computer scientists, engineers and social scientists from around the world, who believed that data and learning can be the best medicine for population health. During the pandemic, they came together to contribute their respective skills and interdisciplinary perspectives to assist in the public health fight against COVID-19 in real time. We therefore hope our work will inspire others to reflect upon the vast untapped potential of building communities of shared purpose to address challenges faced by healthcare systems around the world with meaningful data, diverse and interdisciplinary perspectives and deep domain expertise - in particular in contexts with comparatively limited resources.

The forecasting value produced by this relatively small-scale and ad-hoc cooperative effort illustrates just one of the ways in which countries with limited resources could leverage existing troves of data to conduct real-time data analyses when responding to future pandemics. As the COVID-19 pandemic has made clear that no single organization or even country was fully prepared, this paper calls on governments, universities, health organizations, and industries to invest in and build upon initiatives that create, nurture, and grow these collaborative communities in preparation for the next global disaster.

## Data Availability

All scripts used for forecasting and production of data are located at: https://github.com/sakethsundar/covid-forecaster. All data used for analysis was obtained from the CDC forecast hub repository at: https://github.com/reichlab/covid19-forecast-hub/tree/master/data-processed.

## VIII. Acknowledgments

Leo Anthony Celi is funded by the National Institute of Health through the NIBIB R01 grant EB017205.

## IX. Conflicts of Interest

Patrick Schwab is an employee and shareholder of GlaxoSmithKline plc.

Jade Z.H. Tan is an employee of the U.S. Department of Health and Human Services (HHS)

## Notes

### Funding Statement

No external funding was received.

## References

1. Dong E, Du H, Gardner L. An interactive web-based dashboard to track COVID-19 in real time. The Lancet Infectious Diseases. 2020;20: 533–534. doi:10.1016/S1473-3099(20)30120-1

2. Killeen BD, Wu JY, Shah K, Zapaishchykova A, Nikutta P, Tamhane A, et al. A County-level Dataset for Informing the United States’ Response to COVID-19. 200400756 [physics, q-bio]. 2020 [cited 27 Sep 2021]. Available: http://arxiv.org/abs/2004.00756

3. Couture V, Dingel JI, Green AE, Handbury J, Williams KR. Measuring Movement and Social Contact with Smartphone Data: A Real-Time Application to COVID-19. National Bureau of Economic Research; 2020 Jul. Report No.: 27560. doi:10.3386/w27560

4. Chiu WA, Ndeffo-Mbah ML. Using test positivity and reported case rates to estimate state-level COVID-19 prevalence and seroprevalence in the United States. PLOS Computational Biology. 2021;17: e1009374. doi:10.1371/journal.pcbi.1009374

5. Chen J, Testa C, Watermab P, Krieger N. Intersectional inequities in COVID-19 mortality by race/ethnicity and education in the United States, January 1, 2020–January 31, 2021. Harvard Center for Population and Development Studies.

6. Miller IF, Becker AD, Grenfell BT, Metcalf CJE. Disease and healthcare burden of COVID-19 in the United States. Nat Med. 2020;26: 1212–1217. doi:10.1038/s41591-020-0952-y

7. Badr HS, D. H, Marshall M, Dong E, Squire MM, Gardner LM. Association between mobility patterns and COVID-19 transmission in the USA: a mathematical modelling study. The Lancet Infectious Diseases. 2020;20: 1247–1254. doi:10.1016/S1473-3099(20)30553-3

8. Gibson GC, Reich NG, Sheldon D. REAL-TIME MECHANISTIC BAYESIAN FORECASTS OF COVID-19 MORTALITY. medRxiv. 2020; 2020.12.22.20248736. doi:10.1101/2020.12.22.20248736

9. Chen T, Guestrin C. XGBoost: A Scalable Tree Boosting System. Proceedings of the 22nd ACM SIGKDD International Conference on Knowledge Discovery and Data Mining. San Francisco, California, USA: Association for Computing Machinery; 2016. pp. 785–794. doi:10.1145/2939672.2939785

10. Lundberg SM, Lee S-I. A Unified Approach to Interpreting Model Predictions. Advances in Neural Information Processing Systems. Curran Associates, Inc.; 2017. Available: https://proceedings.neurips.cc/paper/2017/hash/8a20a8621978632d76c43dfd28b67767-Abstract.html

11. Brooks LC, Farrow DC, Hyun S, Tibshirani RJ, Rosenfeld R. Nonmechanistic forecasts of seasonal influenza with iterative one-week-ahead distributions. PLOS Computational Biology. 2018;14: e1006134. doi:10.1371/journal.pcbi.1006134

12. Funk S, Camacho A, Kucharski AJ, Lowe R, Eggo RM, Edmunds WJ. Assessing the performance of real-time epidemic forecasts: A case study of Ebola in the Western Area region of Sierra Leone, 2014-15. PLOS Computational Biology. 2019;15: e1006785. doi:10.1371/journal.pcbi.1006785

13. Yadav SK, Akhter Y. Statistical Modeling for the Prediction of Infectious Disease Dissemination With Special Reference to COVID-19 Spread. Frontiers in Public Health. 2021;9: 680. doi:10.3389/fpubh.2021.645405

14. Funk S, Camacho A, Kucharski AJ, Eggo RM, Edmunds WJ. Real-time forecasting of infectious disease dynamics with a stochastic semi-mechanistic model. Epidemics. 2018;22: 56–61. doi:10.1016/j.epidem.2016.11.003

15. Lagergren JH, Reeder A, Hamilton F, Smith RC, Flores K. Forecasting and Uncertainty Quantification Using a Hybrid of Mechanistic and Non-mechanistic Models for an Age-Structured Population Model. Bulletin of mathematical biology. 2018. doi:10.1007/s11538-018-0421-7

16. Harris CR, Millman KJ, van der Walt SJ, Gommers R, Virtanen P, Cournapeau D, et al. Array programming with NumPy. Nature. 2020;585: 357–362. doi:10.1038/s41586-020-2649-2

17. McKinney W. Data Structures for Statistical Computing in Python. Proceedings of the 9th Python in Science Conference. 2010; 56–61. doi:10.25080/Majora-92bf1922-00a

18. Pedregosa F, Varoquaux G, Gramfort A, Michel V, Thirion B, Grisel O, et al. Scikit-learn: Machine Learning in Python. Journal of Machine Learning Research. 2011;12: 2825–2830.

19. Bertozzi AL, Franco E, Mohler G, Short MB, Sledge D. The challenges of modeling and forecasting the spread of COVID-19. PNAS. 2020;117: 16732–16738. doi:10.1073/pnas.2006520117

20. Cramer EY, Ray EL, Lopez VK, Bracher J, Brennen A, Rivadeneira AJC, et al. Evaluation of individual and ensemble probabilistic forecasts of COVID-19 mortality in the US. 2021 Feb p. 2021.02.03.21250974. Available: https://www.medrxiv.org/content/10.1101/2021.02.03.21250974v1

21. Zou D, Wang L, Xu P, Chen J, Zhang W, Gu Q. Epidemic Model Guided Machine Learning for COVID-19 Forecasts in the United States. 2020 May p. 2020.05.24.20111989. Available: https://www.medrxiv.org/content/10.1101/2020.05.24.20111989v1

22. Pei S, Shaman J. Initial Simulation of SARS-CoV2 Spread and Intervention Effects in the Continental US. 2020 Mar p. 2020.03.21.20040303. Available: https://www.medrxiv.org/content/10.1101/2020.03.21.20040303v2

23. COVIDAnalytics. [cited 27 Sep 2021]. Available: https://www.covidanalytics.io/

24. Khan ZS, Van Bussel F, Hussain F. A predictive model for Covid-19 spread applied to eight US states. 200605955 [physics, q-bio]. 2020 [cited 27 Sep 2021]. Available: http://arxiv.org/abs/2006.05955

25. Chen Y-C, Lu P-E, Chang C-S, Liu T-H. A Time-dependent SIR model for COVID-19 with Undetectable Infected Persons. IEEE Trans Netw Sci Eng. 2020;7: 3279–3294. doi:10.1109/TNSE.2020.3024723

26. Bracher J, Ray EL, Gneiting T, Reich NG. Evaluating epidemic forecasts in an interval format. PLOS Computational Biology. 2021;17: e1008618. doi:10.1371/journal.pcbi.1008618

27. Kandula S, Yamana T, Pei S, Yang W, Morita H, Shaman J. Evaluation of mechanistic and statistical methods in forecasting influenza-like illness. Journal of The Royal Society Interface. 2018;15: 20180174. doi:10.1098/rsif.2018.0174

28. Baker RE, Peña J-M, Jayamohan J, Jérusalem A. Mechanistic models versus machine learning, a fight worth fighting for the biological community? Biology Letters. 2018;14: 20170660. doi:10.1098/rsbl.2017.0660

29. Nicolò C, Périer C, Prague M, MacGrogan G, Saut O, Benzekry S. Machine learning versus mechanistic modeling for prediction of metastatic relapse in breast cancer. 2019 May p. 634428. Available: https://www.biorxiv.org/content/10.1101/634428v1

30. Schwab P, Mehrjou A, Parbhoo S, Celi LA, Hetzel J, Hofer M, et al. Real-time prediction of COVID-19 related mortality using electronic health records. Nat Commun. 2021;12: 1058. doi:10.1038/s41467-020-20816-7

31. Schwab P, Schütte AD, Dietz B, Bauer S. Clinical Predictive Models for COVID-19: Systematic Study. Journal of Medical Internet Research. 2020;22: e21439. doi:10.2196/21439

